# Low-Frequency Dual Target Deep Brain Stimulation May Relieve Parkinsonian Symptoms

**DOI:** 10.1101/2025.03.25.25324612

**Authors:** Rocio Rodriguez Capilla, Aislinn M. Hurley, Karthik Kumaravelu, Jennifer J. Peters, Hui-Jie Lee, Dennis A. Turner, Warren M. Grill, Stephen L. Schmidt

## Abstract

**Background:** Deep brain stimulation (DBS) reduces the motor symptoms of Parkinson’s disease. The two most common targets are the subthalamic nucleus and the globus pallidus. Dual target deep brain stimulation may better reduce symptoms and minimize side effects, but the optimal parameters of dual target deep brain stimulation and their potential interactions are unknown.

**Objective:** Our purpose was to quantify the frequency response of dual target DBS on bradykinesia and beta oscillations in participants with Parkinson’s disease, and to explore intrahemispheric pulse delays as a means to reduce total energy delivered.

**Methods:** We applied dual target DBS using the Summit RC+S in six participants, varying deep brain stimulation frequency.

**Results:** Dual target DBS at 50 Hz was effective at reducing bradykinesia, whereas increasing deep brain stimulation frequency up to 125 Hz also significantly reduced beta power. This frequency effect on beta power was replicated in a biophysical model. The model suggested that 22 Hz dual target deep brain stimulation, with an intrahemispheric delay of 40 ms, can reduce beta power by 87%.

**Conclusion:** We conclude that dual target DBS at 125 Hz best reduced bradykinesia. However, low frequency DBS with an appropriate intrahemispheric delay could improve symptom relief.

## Introduction

Parkinson’s disease (PD) is the second most prevalent neurodegenerative disease (1). Deep brain stimulation (DBS) of either the subthalamic nucleus (STN) or the globus pallidus (GP) is an effective surgical treatment for PD (2–5). Combined DBS of both STN and GP (dual target, DT DBS) may be more effective than DBS of either region alone (6–8). While DT DBS reduced both Unified Parkinson’s Disease Rating Scale (UPDRS) scores and levodopa equivalent daily dose, it remains unknown whether these benefits may be further improved with the optimization of stimulation parameters.

The most widely reported biomarker for PD is the power of beta band (13 – 30 Hz) oscillations in the basal ganglia, which increase prominently in the pathological state (9,10), and there is a correlation between the reduction in beta power in the STN and the improvement in bradykinesia with treatment (8,9,11). We observed a correlation between bradykinesia and STN beta power at different amplitudes of 125 Hz DT DBS (8). Here, we investigate this relationship across DBS stimulation frequencies to quantify the effect of frequency and assess the generalizability of beta as a biomarker for akinetic rigid symptoms across DBS parameters.

Stimulation amplitude and pulse width are determined based on the patient’s tolerance (i.e., side effects) and symptomatic relief (8,12–15). However, the frequency is typically set to 130 Hz, with few changes after the first clinical visit (16,17). Further, studies of DBS frequency focused on differences between a single low frequency (40 – 80 Hz) and a single high frequency (> 100 Hz) (17–19). As well, most reports of frequency differences focused on gait and posture rather than bradykinesia, and multiple studies favor low-frequency DBS, which shows a greater improvement of gait symptoms with fewer side effects (20–23). Conversely, preclinical studies of frequency effects suggest that the response to DBS exists across a spectrum rather than a binary distribution (24,25). There has been no investigation of the effects of frequency on patients with bilateral, DT DBS, and it remains unknown whether parameters appropriate for single target (ST) DBS are also suitable for DT DBS. Here, we quantify the relationship between the frequency of DT DBS and bradykinesia and beta power in human participants with PD and leads implanted bilaterally in the STN and GP. First, we confirm that STN beta power is a suitable biomarker across DT DBS frequencies. We then examine the frequency response of DT DBS over 2 – 125 Hz and compare the effects on beta power to ST DBS.

DT DBS has an additional parameter, the delay between intrahemispheric stimulation pulses in STN and GP. When four stimulation programs are active, the Summit RC+S delivers the pulses with a delay of one-fourth of the total period between pulses, rather than applying pulses simultaneously. Therefore, the delay between pulses is determined by the frequency of stimulation. Since the delay cannot be controlled independently of the stimulation frequency, we used a computational model – widely employed to understand the pathophysiology of PD and the mechanisms of action of DBS (26–29) – to quantify the effects of the delay and the frequency as two independent variables and overcome limitations imposed by available DBS hardware.

## Methods

### Participant selection

Six participants were enrolled and implanted in our clinical trial for PD (NCT #03815656). Due to travel difficulty, one participant (Participant 4) did not perform all of the experiments described in this manuscript (Fig. 1). Inclusion criteria and surgical methods are found in Mitchell et al, 2022 (6). Briefly, leads were implanted bilaterally in both the STN and GP, which were connected to a single Summit RC+S (Medtronic, Minneapolis, MN) device. All participants provided written informed consent. Protocols were approved by the FDA (IDE # G180280), Medtronic External Research Program Board, and Duke University Health System Institutional Review Board.

**Figure 1.**
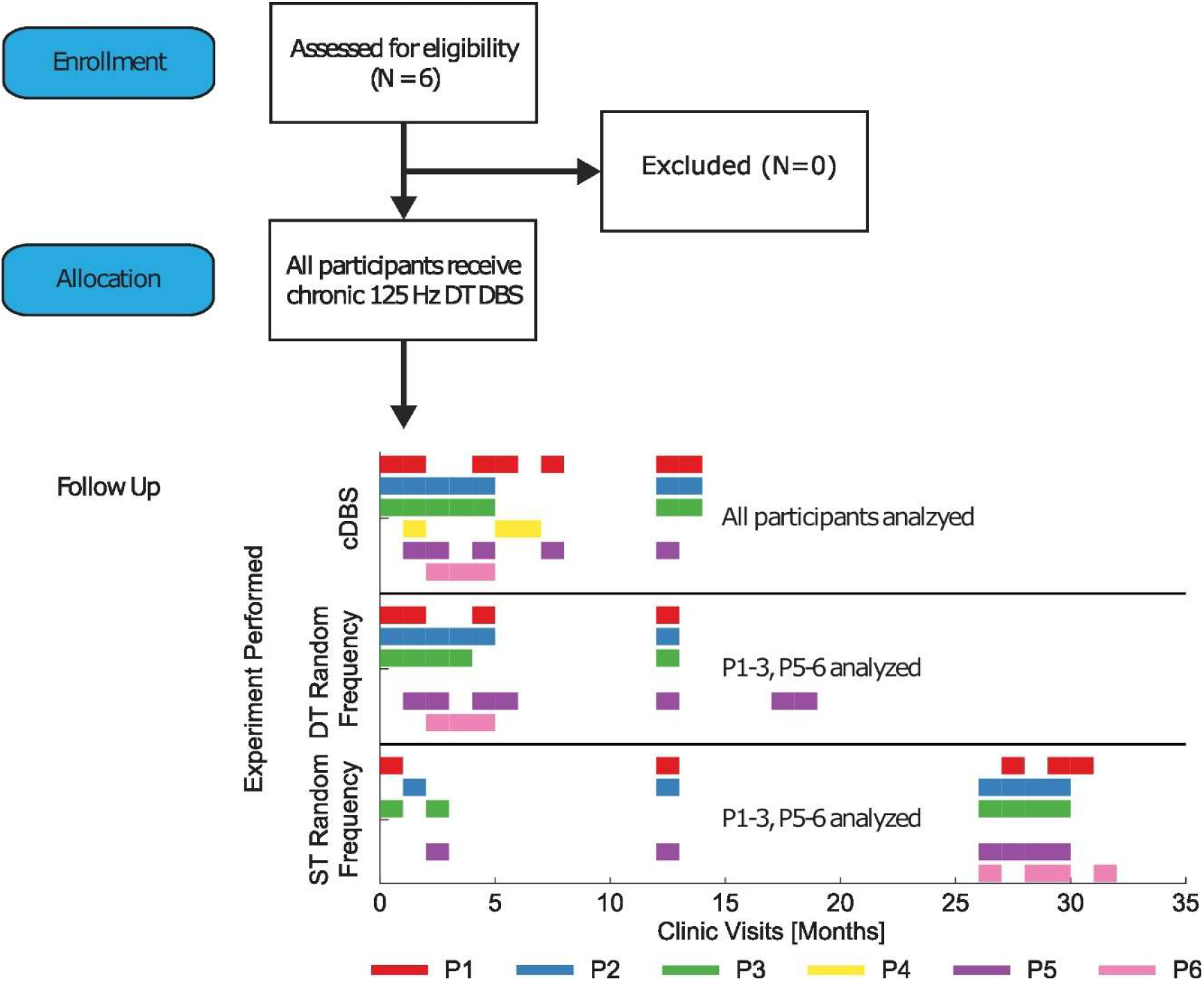
Consort Diagram. Six people with PD consented and were assessed for eligibility. No participant was excluded. All participants received 125 Hz DT DBS between monthly follow-up visits. During monthly follow-up visits, a variety of experiments were conducted. Participant 4 did not perform random frequency experiments. Data from each experiment type was analyzed for all participants who performed the experiments. DT: Dual-target. ST: Single-target (STN only for P1-5, GP only for P6).

### Clinical experimental design

The participants returned to the clinic monthly to participate in a variety of novel experiments. Their implantable pulse generator was connected to the research computer via Bluetooth™. The experiments were completed using a custom-written C# (Microsoft, Redmond, WA) control application that allows manual and computer-generated (e.g., randomized) adjustment of the DBS parameters. Two different experimental paradigms were conducted with human participants: continuous DBS experiments with bradykinesia assessments and random frequency experiments without bradykinesia assessments. The first experiments consisted of paired DT DBS off and DT DBS on trials (Fig. S1A). After approximately 210 s of the off-period, the participants were asked to perform grasping motions with their hands positioned above a 3D infrared camera (Leap Motion Controller, Ultraleap, Mountain View, CA) for a total of approximately 10 s (Fig. S1B). Directly after the completion of the DBS off trial, the frequency was set to 50, 75, 100 or 125 Hz (other parameters were unchanged from best monopolar settings) and DBS was turned on for another 300 s trial. After 210 s, the participants performed the hand-grasp task for another 10 s.

During random frequency experiments to assess beta oscillations, the DBS frequency was randomly changed every 2, 4, or 10 s, depending on the trial, from a pool of 11 frequencies evenly distributed between 2 – 125 Hz (Fig. S2). The other DBS parameters remained unchanged. During this time, participants were seated in a comfortable chair and not directed to perform any task. Only one epoch length of stimulation was applied within each trial. Each trial consisted of only DT DBS epochs or ST DBS epochs. For the trials in which we applied ST DBS, Participants 1 – 5 received STN DBS, while Participant 6 received GP DBS based on participants’ preferences. Each trial lasted for a maximum of 1800 s, however, trials were stopped early if participants reported being too uncomfortable to continue. Any experiment with a duration of less than 300 s was discarded from the final analysis.

### Signal processing

Data analysis of LFP, bradykinesia, and acceleration was performed using custom-written MATLAB (MathWorks, Natick, MA) scripts. Within each 300 s DBS trial, bradykinesia was assessed for 10 s as in Schmidt et al., 2024 (8). The duration of the assessment were manually isolated using the grasp distance output of the Leap Motion (Fig. S1B). Average hand grasp speed was determined by the average peak-to-peak time of grasps completed within the epoch. The beta power of each LFP channel was calculated using the pwelch() function during the duration of the bradykinesia assessments and was integrated over the beta band using the trapz() function. The Pearson correlation coefficients were calculated to between hand grasp speed and beta power (on a decibel scale) in the contralateral STN. The most correlated hand for each participant was determined to be the “best hand”/ “best side” for the random frequency experiments.

For random frequency experiments, the DBS stimulation artifacts were removed from the LFP data using the PARRM method for each epoch independently (Fig. S3) (30). The beta power was calculated with the pwelch() function as it was for the previous experiments. For visualization (but not statistical analysis), observations of beta power were normalized by the median beta power of 125 Hz during DT DBS, per participant. For each participant, the contralateral hemisphere of their best hand as determined during the DBS experiments was used as their “best side” (Table S1).

### Statistical analyses

Statistical analyses were performed in R 4.3.2 (R Core Team, Vienna, Austria) and SAS 9.4 (SAS Institute, Cary, NC). For the cDBS experiments, two random intercept linear models were fitted by using either grasp speed or beta power as the outcome variables with the restricted maximum likelihood (REML) method. The models included DBS frequency (DBS off, 50 Hz, 75 Hz, 100 Hz, or 125 Hz), medication (on/off), the interaction between stimulation frequency and medication, and allowing each participant to vary in the mean response. Because the interaction effect of DBS frequency and medication was not statistically significant, the final model included DBS frequency and medication as covariates. As sensitivity analyses, the above models were repeated using generalized estimating equations.

In random frequency experiments, the primary analysis used data from the best recording site. To assess the association between DBS frequency and beta power, we first determined the functional form of the DBS frequency in relation to beta power using a preliminary random intercept linear model with stimulation frequency as the only covariate, representing DBS frequency as restricted cubic splines with the number of knots varying from 3 to 5. The number of knots was selected from the model with the lowest Akaike information criterion. After the number of knots was selected, we then tested whether the relationship between stimulation frequency and beta power was linear. Because there was evidence of a non-linear relationship between stimulation frequency and beta power, we represent stimulation frequency as restricted cubic splines. Next, a random intercept linear model was estimated using the REML method by including stimulation frequency (using restricted cubic splines), DBS configuration (ST DBS, or DT DBS), epoch length (2, 4, 10 s), the interaction between DBS frequency and DBS configuration as covariates, and allowing each participant to vary in the mean response. Two sensitivity analyses were conducted: First, the above analyses were repeated using data from both the best recording side and the contralateral side. Second, a linear generalized estimating equations model with robust error estimation was fit for the best side using the same model specification from the primary analysis, which relaxed the normally distributed residuals assumption.

### Computational model and experiment design

We adapted the previously published biophysical computational model of the rodent cortico-basal ganglia-thalamic closed-loop (26). The original model featured 10 biophysical neurons in each of the following regions: the cortex (CTX), the striatum, STN, globus pallidus externa (GPe), GPi, and thalamus (Th). We modified the model to include GP DBS and DT DBS. We then adapted the model to human physiology and validated its response to phase-targeted DBS (Supplemental Material, Table S2-S4).

To analyze the frequency response, the DBS frequency was varied from 0 – 200 Hz in steps of 5 Hz in independent simulations. The beta power was calculated, and the median beta power across 10 trials per frequency was plotted in a frequency tuning curve. The same process was used for the analysis of ST and DT DBS.

To conduct intertarget delay simulations, we applied cDBS and varied the intertarget pulse delay as a new parameter. The delay was introduced as the duration (in ms) from the rising edge of the STN pulse to the rising edge of the GPi pulse. Across iterations, delays were swept from 0 to 46 ms with 2 ms steps. We removed the DBS artifact by blanking during the time of stimulation (0.1 ms before to 2 ms after), calculated beta power using the pwelch() function, and then integrated over the beta band (13 – 30 Hz) using the trapz() function. For display, beta power was normalized by dividing by the beta power at baseline (PD condition, without DBS).

## Results

### Effects of continuous DBS

We quantified the effects of DT DBS at different frequencies on beta power and hand grasp speed using random intercept linear models (Fig. S1).The interaction effect of frequency and medication on grasp speed was not statistically significant (p = 0.51). Therefore, the final statistical model included DBS frequency and medication only, not their interaction. DBS at any frequency (50 Hz, 75 Hz, 100 Hz, or 125 Hz) increased grasp speed relative to DBS off (Table S5), and on medication increased grasp speed by 0.13 Hz (95% CI 0.03 to 0.23 Hz, p = 0.01) compared to off medication. However, we did not observe a significant difference in grasp speed when increasing stimulation frequency by 25 Hz from 50 Hz to 125 Hz (Fig. 2A, S4).

**Figure 2.**
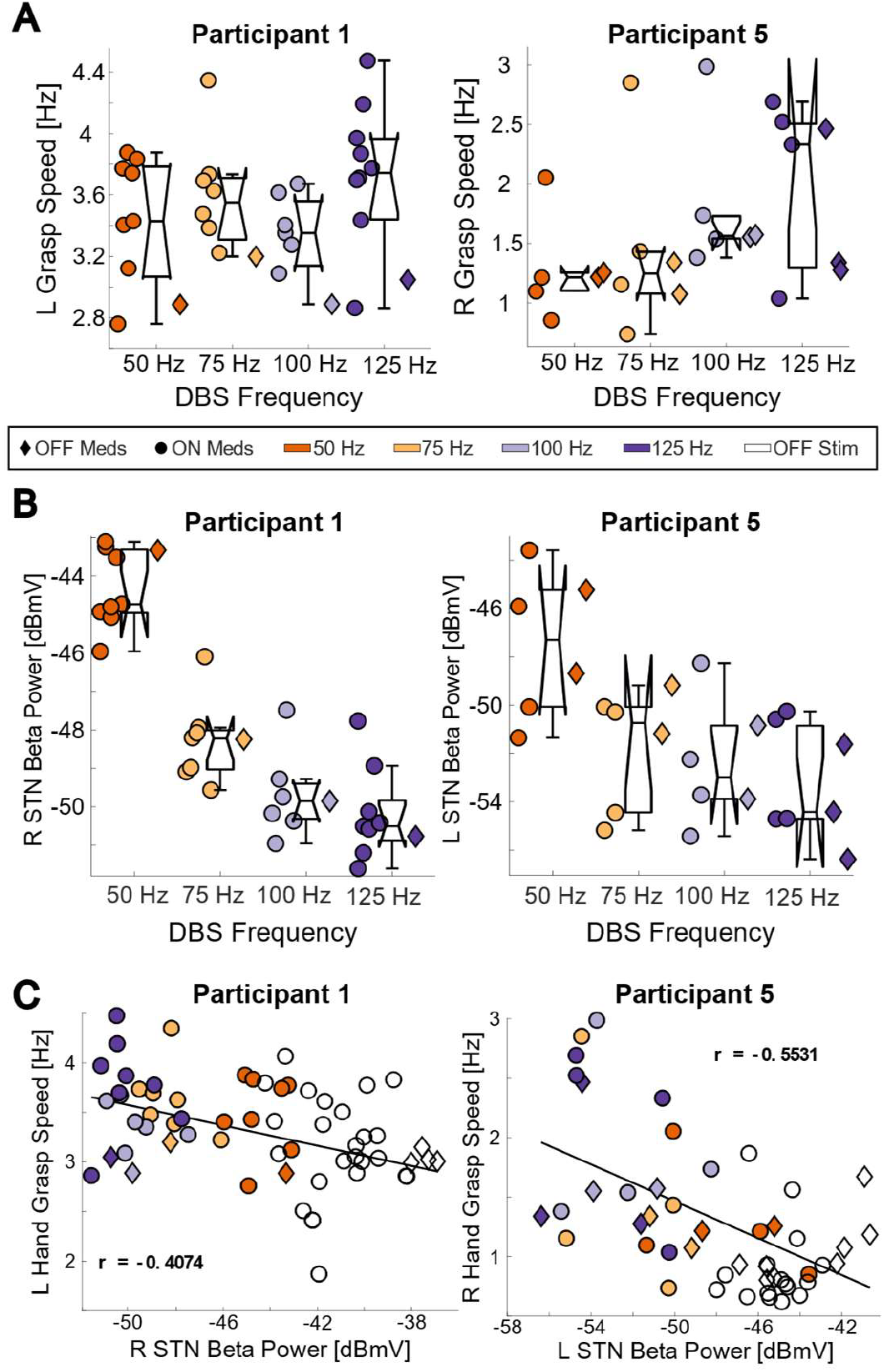
DT DBS reduced bradykinesia and beta power. **A)** Hand grasp speed as a function of stimulation frequency. Median grasp speed increased at 125 Hz DT DBS for Participants 1 & 5. Data for all participants are included in Fig. S4. **B)** Beta power as a function of stimulation frequency. For both participants, beta power decreased with increasing DBS frequency. See Fig. S5 for all participants. **C)** Scatter plots of hand grasp speed versus contralateral STN beta power. Trials with lower grasp speeds (i.e. more bradykinesia) corresponded to higher beta power. Black lines represent best-fit lines. See Fig. S6 for all participants. r Pearson’s correlation. L left. R right.

We then considered the effect of DBS on beta power. The interaction between DBS frequency and medication on beta power was not significant (p = 0.35). We observed DBS at 50 Hz, 75 Hz, 100 Hz, or 125 Hz DBS reduced the mean beta power relative to no DBS, and 125 Hz DBS reduced beta power more than any other setting (e.g., −0.84 dB compared to 100 Hz, 95% CI −1.63 to −0.06 dB, p=0.03; Fig. 2B, S5). We observed a numerical difference in mean beta power between on and off medication that was not significant (mean difference: −0.39 dB, 95% CI −0.81 to 0.04 dB, p = 0.08).

To assess the suitability of beta power as a biomarker for bradykinesia within the cohort, we quantified the Pearson’s correlation between grasp speed and beta power per participant. Four of the six participants exhibited a significant negative correlation (−0.553 ≥ *r* ≥ −0.136) between grasp speed and beta power in the contralateral STN (Fig. 2C, S6, S7). Of the two participants without significant correlation, one of them contributed limited data (Participant 4), and the other (Participant 6) exhibited limited bradykinesia that did not respond to DBS, and therefore no correlation was expected.

### Random frequency DBS experiments

To investigate further the effect of DBS frequency on beta power in the STN, we rapidly switched DBS frequencies randomly over the range 2 – 125 Hz by 12.3 Hz intervals during both DT DBS and ST DBS (Fig. S2). Beta power decreased with increasing DBS frequency for both DT and ST DBS (Fig. 3A, B). DT and ST DBS produced similar patterns across frequencies, and beta power decreased more during the use of DT DBS than during ST DBS with the largest difference observed at 38.9 Hz (Fig. 3C). The beta power in each epoch was not dependent on the DBS frequency of the previous epoch (see Supplemental Results, Fig. S8). Using a random intercept linear model with restricted cubic splines for stimulation frequency, increasing the frequency by 12.3 Hz led to a significant decrease of the mean beta power in a range of 0.1 to 2 dB, depending on frequencies, for both DT and ST DBS (Table S6). The largest mean decrease in beta power was observed when the frequency was increased from 38.9 to 51.2 Hz, where the mean beta power decreased by 2.023 dB (95% CI −2.124 to −1.922 dB, p < 0.001) and by 1.634 dB (95% CI −1.719 to −1.549 dB, p < 0.001) for DT and ST DBS, respectively. While the mean beta power across participants showed a reduction from 14.3 to 26.6 Hz, the per-participant results show a numerical increase in beta power at that frequency in 6 out of the 10 STNs (for P5 and P6 on the best side, P1, P2, P3 and P6 on the contralateral side, Fig. 3A).

**Figure 3.**
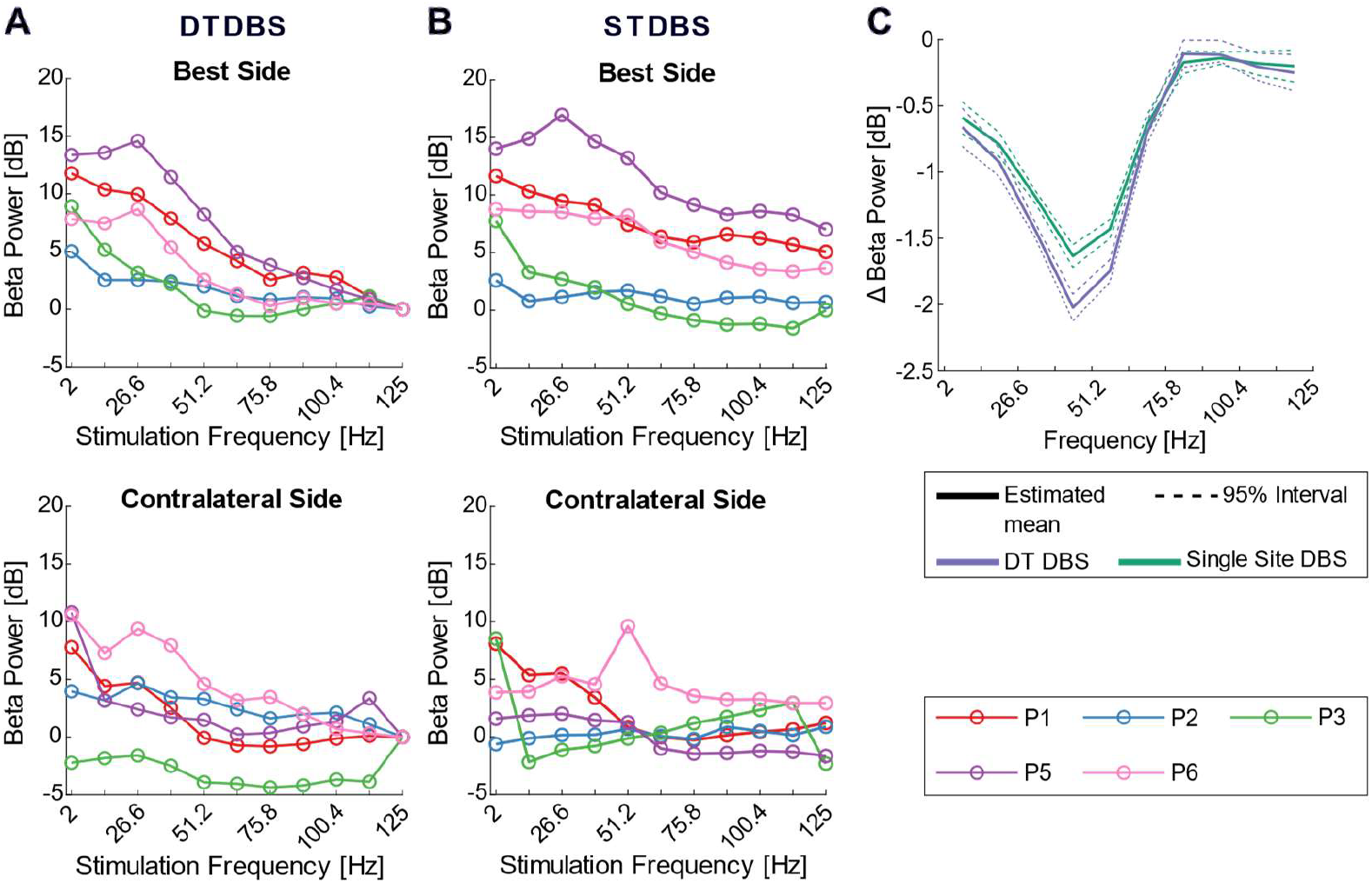
Beta power decreased with increasing frequency of DT and ST DBS. Median beta power (normalized to 125 Hz DT DBS) as a function of stimulation frequency for DT DBS (**A**) and ST (STN for P1, P2, P3 and P5, or GP for P6) DBS (**B**) for the hemisphere most correlated with bradykinesia (top) and the contralateral hemisphere (bottom). DT DBS reduced beta power to a greater extent than ST DBS. Participant 4 did not perform these experiments. **C)** Mean difference (solid line) in beta power for each increase in DBS frequency for DT DBS (lilac) and ST DBS (green) and 95% interval (dotted lines).

### Computational model predicts effective low frequency DBS

We used a biophysically based computational model of the human motor cortico-basal ganglia-thalamo-cortical loop (Fig. 4A) to quantify the effects of DBS frequency and intertarget delay beyond what is possible with currently implanted devices. After validating the model (Supplemental Material, Fig. S9), we generated frequency tuning curves for both DT and STN DBS (Fig. 4B–D). Generally, beta power decreased with increasing DBS frequency for both DT and STN DBS, and beta power reached minima at 105 Hz (DT DBS) and 110 Hz (STN DBS). Beyond these minima, increasing the frequency led to slight increases in beta power. DT DBS produced a greater reduction in beta power than STN DBS, but the difference was slight.

**Figure 4.**
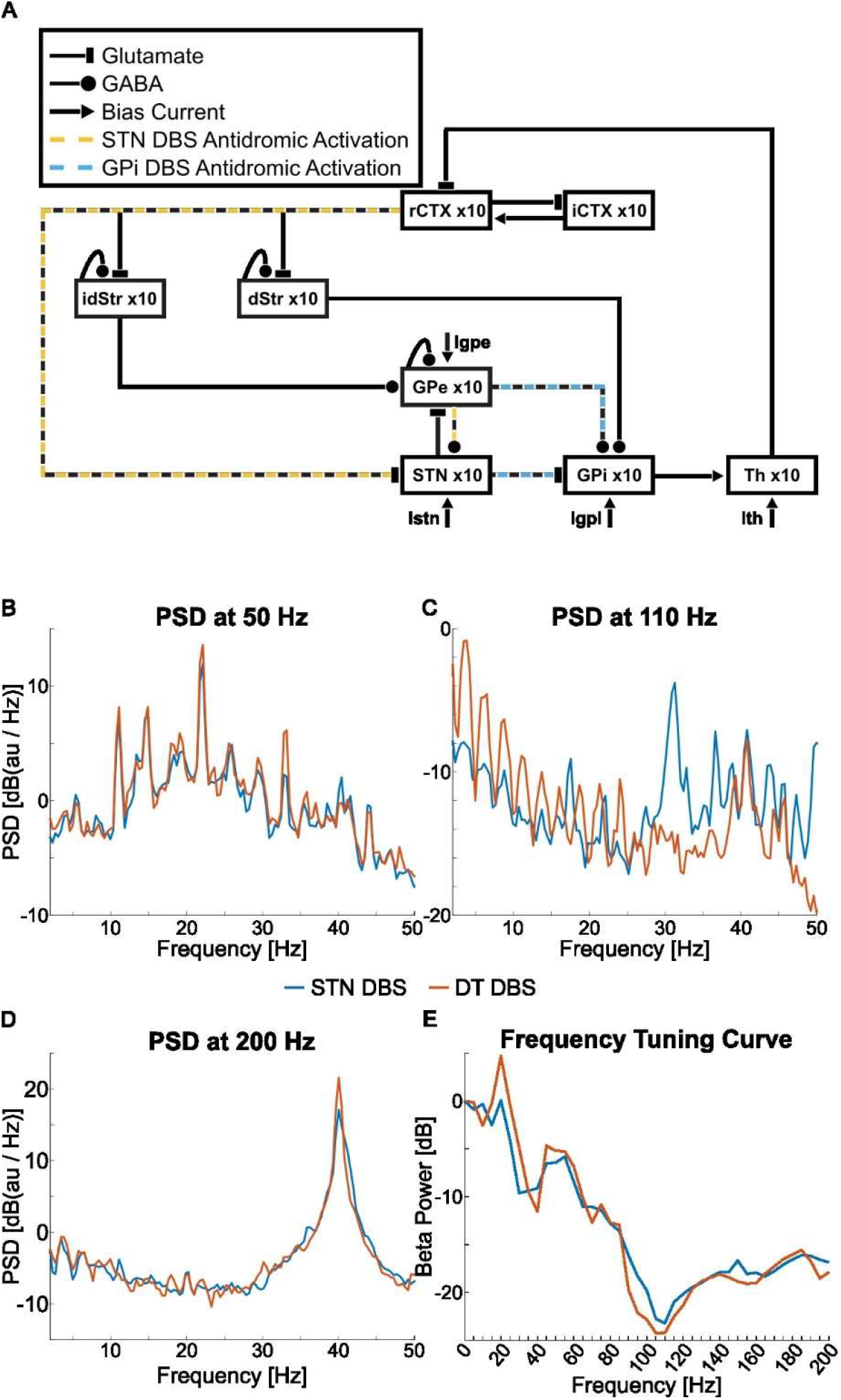
Schematic of model and effect of DBS frequency on beta power in the computational model. A). Model topology, including the number of neurons per area of the brain modeled and interconnections. The color indicates antidromic activation due to DBS of the STN (yellow) and the GPi (blue). Bias currents (I) were applied to the STN, the GPi, the GPe, and the Th to set baseline firing rates. Abbreviations: (r/i)CTX Cortex, (id/d)Str Striatum, GP(e/i) Globus pallidus (externa/interna), STN Subthalamic nucleus, Th Thalamus. B-D)DT DBS (red) and STN DBS (blue) from 0 – 200 Hz. A – C) Power Spectral Density (PSD) for key points along the frequency tuning curve. D) Beta power (normalized to DBS off) over 0 – 200 Hz DT and STN DBS.

The model responded appropriately to adaptive DBS based on the phase of the STN beta oscillation (Supplemental Material). Next, we quantified the effects of varying the delay between the DBS pulses delivered to the STN and the pulses delivered to the GPi during open-loop DT DBS. We set the DBS frequency to that of the peak beta power (22 Hz) and changed the intertarget delay from 0 to 46 ms in increments of 2 ms (Fig. 5A). The frequency of DBS was chosen to match the peak of the beta power to entrain most easily the endogenous beta oscillation by excitatory pulses delivered in the STN. The beta power was increased or decreased by DBS depending on the delay between STN and GPi pulses. DBS with intertarget delays greater than 30 ms decreased beta power and reached a minimum at 40 ms (12.1% of baseline, Fig. 5Bi, 5C). When the delay was set between 8 ms and 30 ms, beta power increased compared to baseline, with a maximum of 164% of baseline power (12 ms, Fig. 5Bii, 5C). The dependence of beta power on delay – decreasing with short delays (i.e. 0 – 6 ms), increasing with longer delays (i.e., 8 – 30 ms), and decreasing again with very long delays (> 30 ms) – was replicated across DBS frequencies in the beta range (i.e. 16 Hz and 28 Hz, Fig. S12). However, when the delay was nearly a complete period long (i.e., the GP pulse slightly led the STN pulse), beta power increased. The reduction in beta power was not as large as during high-frequency DT DBS or 130 Hz ST DBS (12.1% vs 1.02% and 1.39% residual beta power respectively), but the DBS frequency was reduced from 125 Hz to 22 Hz thereby reducing the TEED by 82.4% compared to 125 Hz DT DBS and by 66.1% when compared to 130 Hz STN DBS.

**Figure 5.**
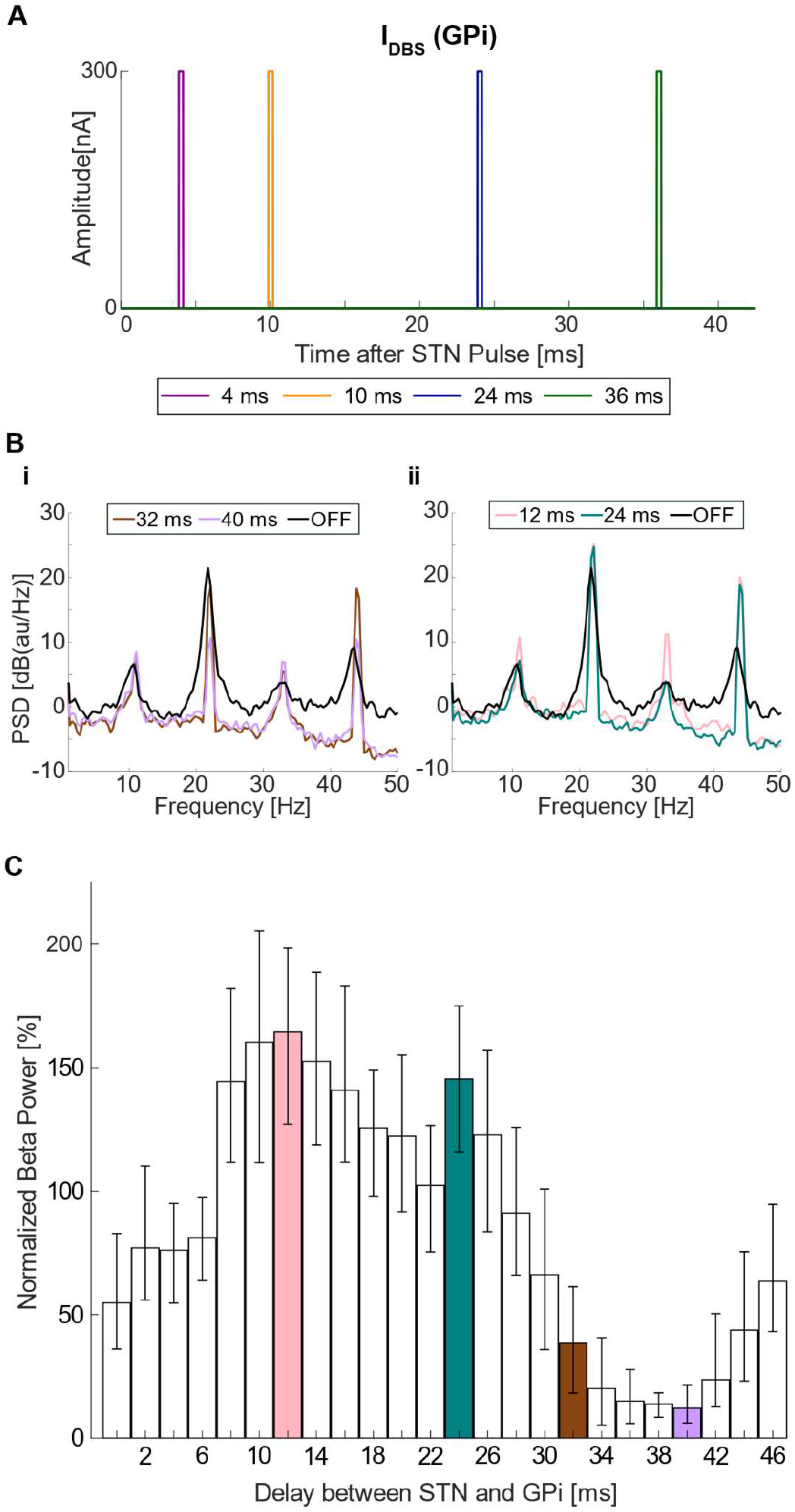
Effect of internuclear pulse delay on beta power. **A)** Example GPi stimulation pulse (STN pulse delivered at time t = 0) across a range of internuclear delays. **B)** PSDs at different delays compared to DBS off (black). **C)** Beta power across delays between the STN and GPi when stimulating at 22 Hz. 100 percent is baseline beta power. Error bars show the range of beta power across 10 trials.

## Discussion

We quantified the relationship between symptoms (bradykinesia), a biomarker (beta power), and DBS frequency in humans and in a computational model. All tested DT DBS frequencies led to significantly higher grasp speeds than no stimulation. This differs from STN DBS which shows limited efficacy at less than 100 Hz (25,31,32). However, we did not observe a significant difference in grasp speed when further increasing DBS frequency beyond 50 Hz. We then conducted experiments with a wider range of frequencies using both ST DBS and DT DBS. Rapid, random switching of DBS frequency revealed that increasing the frequency of DBS by 12.3 Hz led to decreased beta power, a result recapitulated by our biophysical model. DT DBS yielded significantly lower beta power than ST DBS. In the same model, we examined low-frequency DT DBS. We observed that with a 40 ms delay between STN and GPi pulses, DBS delivered at the peak beta frequency in the STN (22 Hz) reduced beta power by more than 88%. In contrast, the actual clinically applied pulse delay between ipsilateral STN and GP with the RC+S at 22 Hz is 11.4 ms, which the model predicted would increase beta power above baseline (Fig. 5C). Indeed, in the human cohort, DT DBS at 26.6 Hz with 9.3 ms intrahemispheric delay increased beta power in 6 of 10 STNs. Additionally, in previous studies, 20 Hz STN DBS increased pallidal and cortical beta oscillatory activity (9,33).

Random frequency experiments highlighted the negative correlation between beta power and DBS frequency and also revealed that DT DBS reduced beta power more than ST DBS. We previously observed a greater reduction in beta power with DT DBS than either STN or GPi DBS alone during UPDRS-III testing (6) in this cohort, and this aligns with greater overall symptom reduction with DT DBS observed in our participants (8). However, there was large inter-participant variability. While beta power for Participant 5 was strongly linearly correlated with DBS frequency, beta power in Participant 2 exhibited much less frequency dependence (Fig. 3A). Variability in bradykinesia reduction with DBS across participants has been observed before (34). Nonetheless, DT DBS frequency and beta power remained significantly correlated across the cohort. In the literature, there is a discrepancy in the effect of DBS frequency on bradykinesia, with some favoring high-frequency DBS (23,35,36). Conversely, other studies did not observe significant differences between frequency settings (20). Most of these studies, however, examined symptoms only and did not include STN beta power. While DT DBS appears promising in initial reports (7,8), larger cohorts are required to validate these findings.

Due to the limitations of the current implanted RC+S hardware, we used a validated computational model to quantify the effects of intertarget pulse timing, and more specifically, whether applying DBS at different phases of the beta oscillation in the STN would modulate the effectiveness of DBS (Supplemental Material). Variable timing of pulses relative to the beta phase affected beta power in humans and non-human primates (37–39). In our model, phase-based aDBS always reduced beta power compared to DBS off. However, the timing of the pulse relative to the beta oscillation modulated the extent of this reduction. We then attempted to regulate the timing of the network by stimulating with beta frequency DBS in the STN (22 Hz) and subsequently suppressing the beta oscillations with the GPi pulse by changing the delay between the two ipsilateral targets. The delay implemented by the Summit RC+S (1/4 of the frequency pulse, ~11 ms) produced nearly double the beta power from the DBS off state in the model. This prediction agrees with the increased beta power observed with beta frequency stimulation in the random frequency experiments (Fig 2). However, a 40 ms delay in the model resulted in an 88% reduction in beta power, and similar results were observed at other DBS frequencies. These results suggest that substantially lower DBS frequencies with a programmable (rather than fixed) STN to GP pulse delay could lead to effects similar to high-frequency DBS. If replicated in humans, this parameter change could lead to longer battery life and reduced side effects that result from constant, high-frequency DBS.

Our study has several limitations. We recruited a small cohort of only six participants. The correlation between bradykinesia and beta power was observed in only four of our six participants further reducing the size of some analyses. Herein, we focused on beta power and its correlated symptoms. However, akinetic and rigid symptoms are only a subset of symptoms observed in PD and patients with axial and gait symptoms might benefit from low-frequency DBS (20,21). The Summit RC+S device also has a frequency range of 2 – 125 Hz when applying DT DBS. We were unable to determine the effect of increasing the DBS frequency beyond 125 Hz nor probe the effect of intertarget delay in human participants.

## Acknowledgments

The authors thank the participants and their caregivers for participating in this study. The authors also thank Candace Boyette, Katherine Genty, Jessica Carlson, and Aparna Choudhury for their assistance in collecting the data. The authors thank Brian Young for implementing the code to perform random frequency experiments. The authors thank Bella Santos for her assistance in producing Figure 4.

## Author Roles

Rocio Rodriguez Capilla: Methodology, Formal Analysis, Investigation, Data Curation, Writing – Original Draft, Visualization.

Aislinn M. Hurley: Methodology, Formal Analysis, Investigation, Writing – Review and Editing.

Karthik Kumaravelu: Methodology, Formal Analysis, Investigation, Writing – Review and Editing.

Jennifer J. Peters: Investigation, Resources, Data Curation, Writing – Reviewing and Editing.

Hui-Jie Lee: Formal Analysis, Writing – Review and Editing.

Dennis A. Turner: Conceptualization, Writing – Review and Editing, Funding Acquisition.

Warren M. Grill: Conceptualization, Writing – Review and Editing.

Stephen L. Schmidt: Conceptualization, Methodology, Investigation, Writing – Original Draft, Supervision.

## Data Availability

To preserve the anonymity of the participants, participant data are available upon request and a data use agreement. Computational data may be generated by the provided code (see Code Availability).

## Code Availability

Code for the computational model will be available at https://gitlab.oit.duke.edu/sls133/human-cortex-basal-ganglia-thalamus-network-model upon publication. Code to analyze human data is available upon request and a data use agreement.

