## Supplemental Material for "Low-Frequency Dual Target Deep Brain Stimulation May Relieve Parkinsonian Symptoms"

### Supplemental Methods and Results

#### Random frequency DBS experiments

We sought to examine any outlasting effect of the previous DBS frequency on the next window of LFP. Because the test windows were short (2 to 10 s) there was a possibility of outlasting effect from the previous window (Fig. S8). For all participants, we observed similar values of beta power regardless of the frequency used in the previous window. This demonstrated that at any given time, the DBS frequency from the previous window had little impact on beta power.

#### Adaptation of the original model to the human basal ganglia

Randomization was applied to the initial conditions of the different cell types, conductance, and the stimulated subset of neurons across ten trials per experimental condition. Beta power was calculated from the LFP using pwelch() and integrated over the beta band using trapz(). After incorporating GP DBS into the model, we found that without antidromic activation of the STN, GPi DBS had little effect on the beta power. Further, modeling studies using realistic axonal pathways have shown DBS to antidromically activate regions through afferent axons projecting to the stimulated nucleus (2,3). Therefore, we added antidromic activation within regions connected to the stimulated region (e.g. GPe and CTX in the case of STN DBS, STN and GPe for GPi DBS). Antidromic activation latency was the same as the synaptic delay between the regions. Because complete activation of all ten neurons must override endogenous network activity, DBS and antidromic activation were only applied to 60% of the neurons, chosen at random.

We adapted the model to the human basal ganglia. To do so, we modified the synaptic delays for key regions (Table S2)(4,5) and the antidromic activation. To achieve mean firing rates found in primates (6,7), we altered the current bias in the GPi, GPe, and Th. We also added a bias current injected directly into the STN. The magnitude of the bias currents was determined empirically with a parameter sweep and was evaluated by minimizing the difference in mean firing rate observed in literature across the STN and GPi/e (Table S3 and S4). The LFP of the STN, GPi, and GPe were calculated by summing the transmembrane potentials of all cells in a region at each time point (1).

To produce average firing rates observed in the literature, we conducted a parameter sweep of the bias currents of the STN, GPi, GPe, and Th. We searched for the best parameters by summation of the difference between the mean firing rates (MFR) values found in the literature and the MFRs from the model in the STN, GPi, and GPe (6,7)(Table S4). The final mismatch from the target was 3.10 action potentials per second.

#### Model validation

With the bias currents in place, we examined the PSD for the healthy state, the PD state without DBS, and the PD state with 130 Hz DBS (Fig. S9A). We observed an increase in oscillatory activity in the beta band, with peak activity at 22 Hz in the PD state compared to the healthy condition. When DBS was applied, the oscillatory activity was reduced. 130Hz DBS reduced the oscillations in the beta band within the model, as observed in humans with PD.

We analyzed the response of the globus pallidus neurons to cortical stimulation, a supra-threshold stimulus pulse was applied to each cortical neuron in the PD state. We quantified the peristimulus time histograms (PSTH) of the GPi and the GPe. The analysis for the PSTH started at 50 ms prior to the cortical stimulus and ended 350 ms after. The final plot was averaged over 100 trials of 10 neurons with a bin width of 1 ms. We compared the model results (Fig. S9B) to those previously observed in humans with PD. We observed a similar response from the model as observed in Nishibayashi et al, 2011, (their Figure 1A2, A4 and A6) (8). In the GPi, a peak activity period was followed by a protracted period of inhibition and another peak in activity (Fig. S9Bi). In the GPe, we observed a short period of inhibition, followed by a peak in activity, before resuming regular behavior (Fig. S9Bii).

#### Adaptive DBS

To test the response of the model to phasic stimulation, we designed a controller based on Escobar Sanabria et al (9)(Fig. S10A). The aDBS controller assessed the phase of the LFP in the STN once every 1.3 ms. To calculate the phase, we first applied a 6 Hz wide second-order non-causal Butterworth filter centered around the peak frequency of the beta oscillation to the STN LFP signal. The instantaneous phase of the oscillations was calculated with a Hilbert transform and the MATLAB function angle() through the last 10 ms. However, the phases from the last 0.5 ms were excluded to account for the settling time of the filter. We tested the controller at multiple phases for the application of DBS—each condition consisted of a range of π/12 between - π and π. If the phase of the oscillation calculated fell within the range, a stimulation pulse (300 nA with a pulse width of 0.3 ms) was applied to the GPi (with antidromic activation). The controller was then disabled for the following 20 ms allowing a maximum DBS frequency of 50 Hz. We performed ten iterations of 10 s simulations with each condition.

We then assessed if the controller was activated within the correct window. For each controller setting, DBS was delivered only if the phase was within the boundaries determined and not anytime else (Fig. S10). To calculate the difference in total electrical energy (TEED) delivered during the aDBS experiments, we determined that DBS amplitude, DBS pulse width, and contact impedance would not change compared to cDBS. We therefore determined the maximum average pulse rate for the simulation that most reduced beta power (i.e. π/6 to π/4); the equivalent DBS frequency was 27.60 Hz. The calculated TEED (10) was 21.23%. Thus, using aDBS reduced TEED by 78.77% compared to the standard 130 Hz GP DBS.

To analyze this data, we removed the DBS artifact by blanking during the time of stimulation (0.1 ms before to 2 ms after), calculated beta power using the pwelch() function, and then integrated over the peak beta band (19 – 25 Hz) using the trapz() function. For display, beta power was normalized by dividing by the beta power at baseline (PD condition, without DBS).

While a decrease in beta power was observed at all different settings of the controller, there were stark differences in beta power depending on the oscillation phase when stimulation was delivered (Fig. S11). Pulses delivered to the GPi during STN beta oscillation phases between -3π/4 and -7π/12 reduced beta power to about 70% of baseline power (or a reduction of ~1.5 dB). However, when the controller applied pulses between π/6 and π/3, beta power was reduced to less than 20% of the baseline beta power (80% or ~7.0 dB reduction) while maintaining a lower total electrical energy delivered (TEED) compared to 130 Hz ST DBS.

#### Intrahemispheric delay experiments

By varying the delay of pulses between the STN and GPi, we observed a large decrease in beta power when DBS was matched to the frequency of the ongoing oscillation. However, it was unclear whether it was necessary to match the frequency of the endogenous beta oscillation. Therefore, we repeated latency sweep simulations with DBS frequencies mismatched to the frequency of the beta oscillation, both 16 Hz and 28 Hz DBS. The period of the DBS limited the range of latencies tested. Intrahemispheric latency could both potentiate or reduce the beta band oscillation with DBS mismatched to the endogenous beta oscillation (Fig. S12). Indeed for 16 Hz the overall shape of the change with increasing latency matched that resulting from 22 Hz DBS. However, during 22 Hz DBS the reduction in beta power was greater (i.e. 87.9%) compared to either 16 Hz or 28 Hz DBS (a reduction of 80.0% and 78.0%, respectively).

### Supplemental Figures

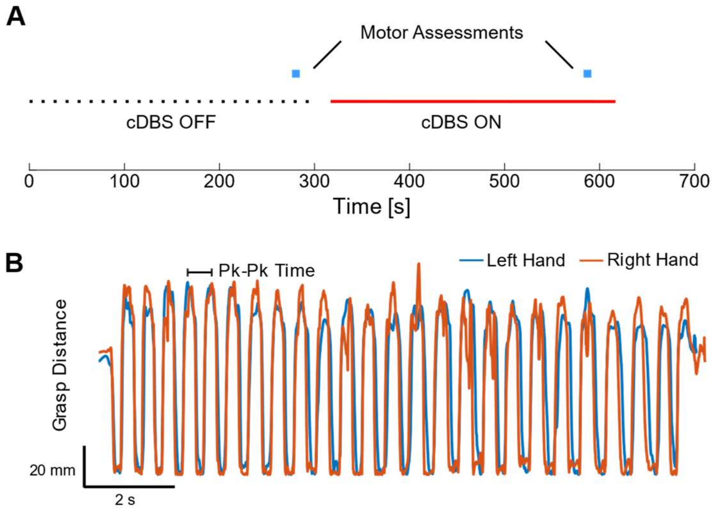


**Figure S1. Experimental design of DBS experiments to measure the effects of DBS frequency on bradykinesia and beta power in humans with Parkinson's disease. A)** Timing for experiments. The experiment consisted of two consecutive trials: during the first trial DBS was not applied (DBS off), and during the second DBS was delivered at 50, 75, 100, or 125 Hz (DBS on). Bradykinesia was assessed for 10 s near the end of each trial. **B)** During the measurement of bradykinesia, participants were asked to open and close their hands as rapidly as possible. The average speed was calculated using the positive peak-to-peak time for each grasp and averaged across all grasps.

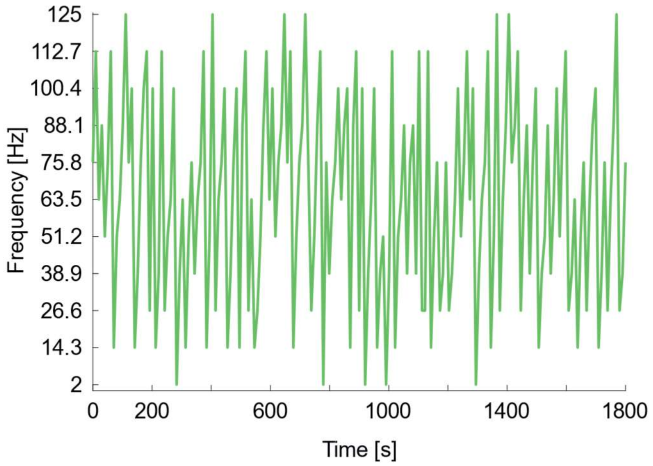


**Figure S2. Experimental design for random frequency experiments.** DBS frequency was changed every 2, 4, or 10 s for a maximum duration of 1800 s. The selected frequency was randomized between 11 frequencies that were evenly distributed between 2 and 125 Hz were tested in total.

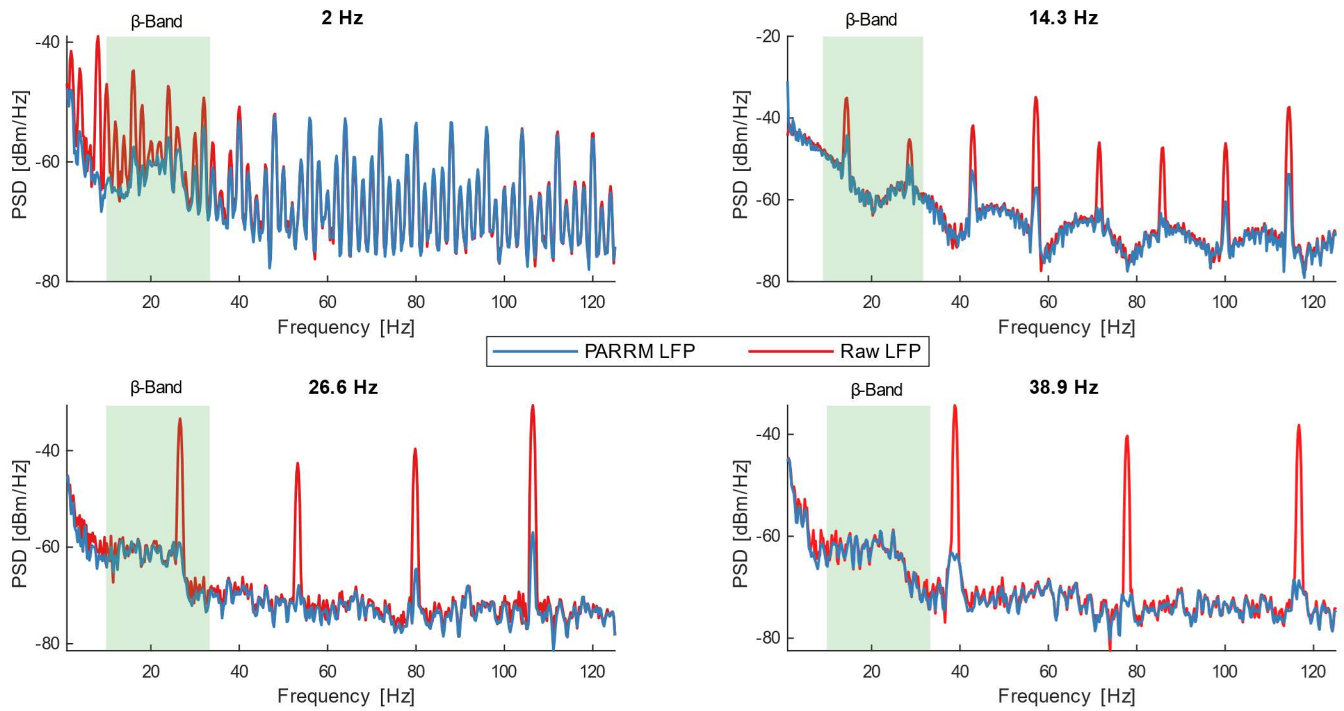


**Figure S3. Removal of simulation artifact on beta band frequencies.** Depending on the DBS frequency beta power could be obscured by stimulation artifacts (red lines). To mitigate the DBS artifact, the PARRM algorithm was applied before calculating the PSD (blue lines). PARRM greatly reduced but did not completely remove DBS artifacts in all cases.

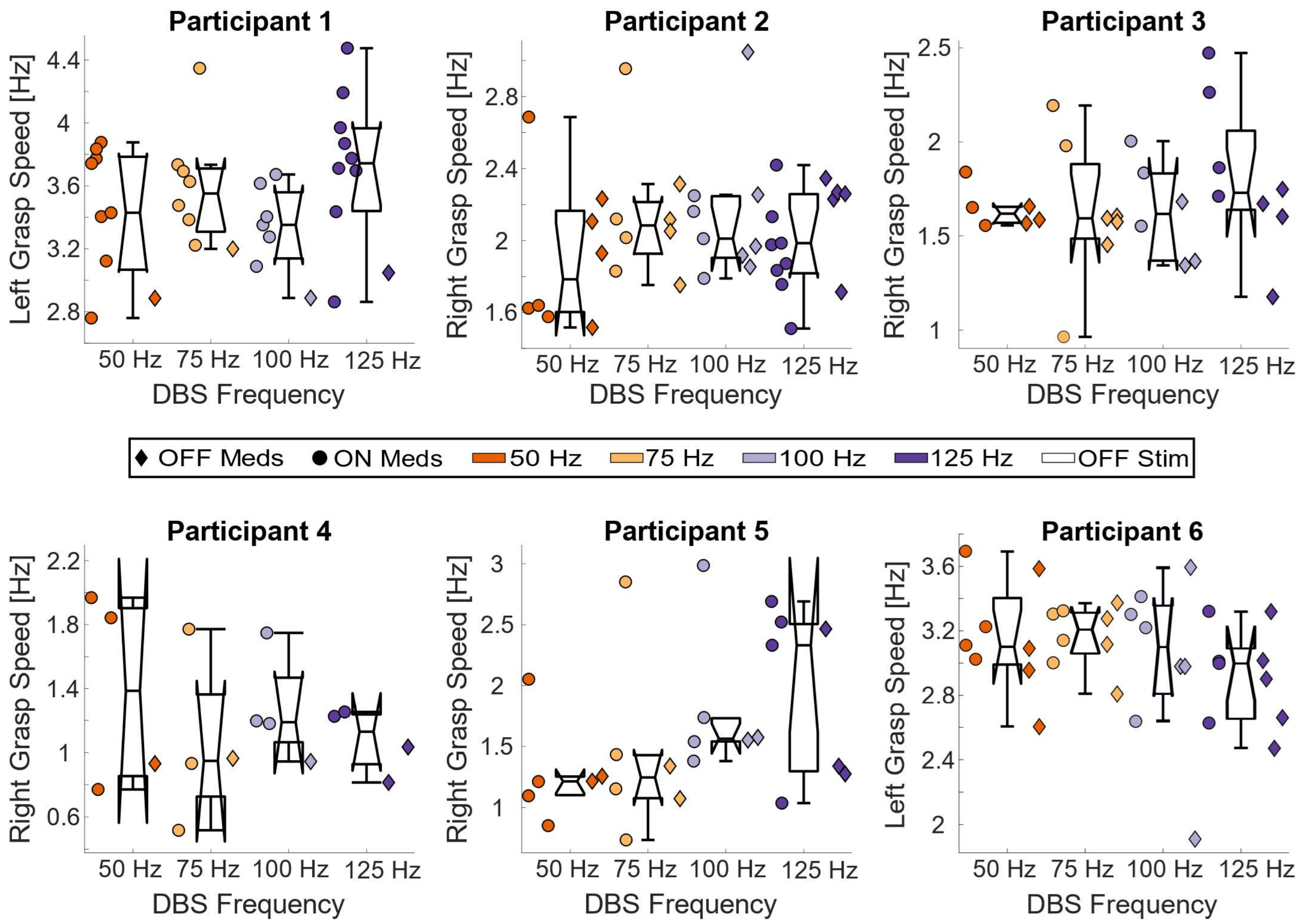


**Figure S4. Grasp Speed per experiment type per participant.** Grasp speeds for each experiment for each participant.

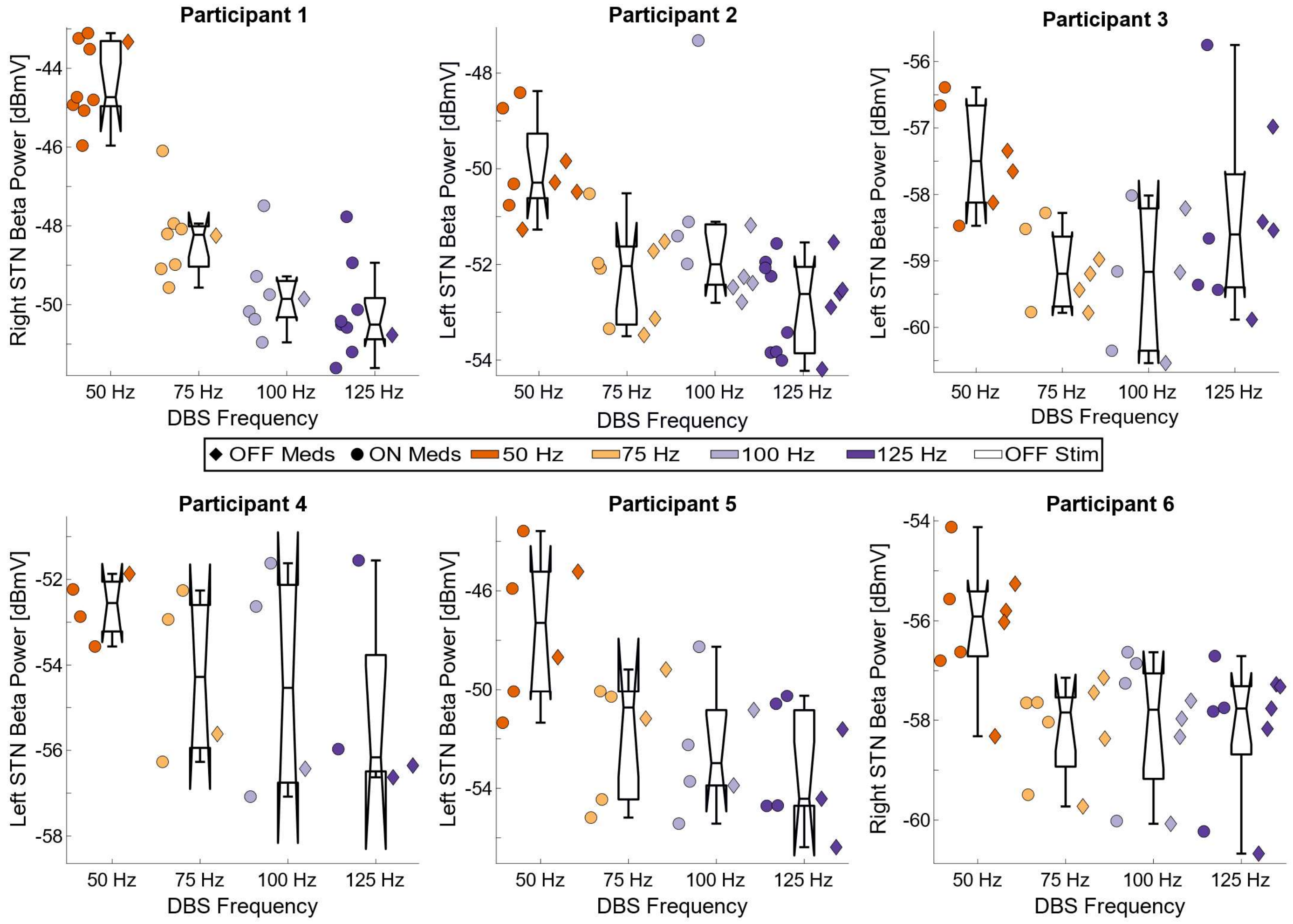


**Figure S5. Beta power per experiment type per participant.** Beta power in the contralateral STN during movement of the best hand for each participant.

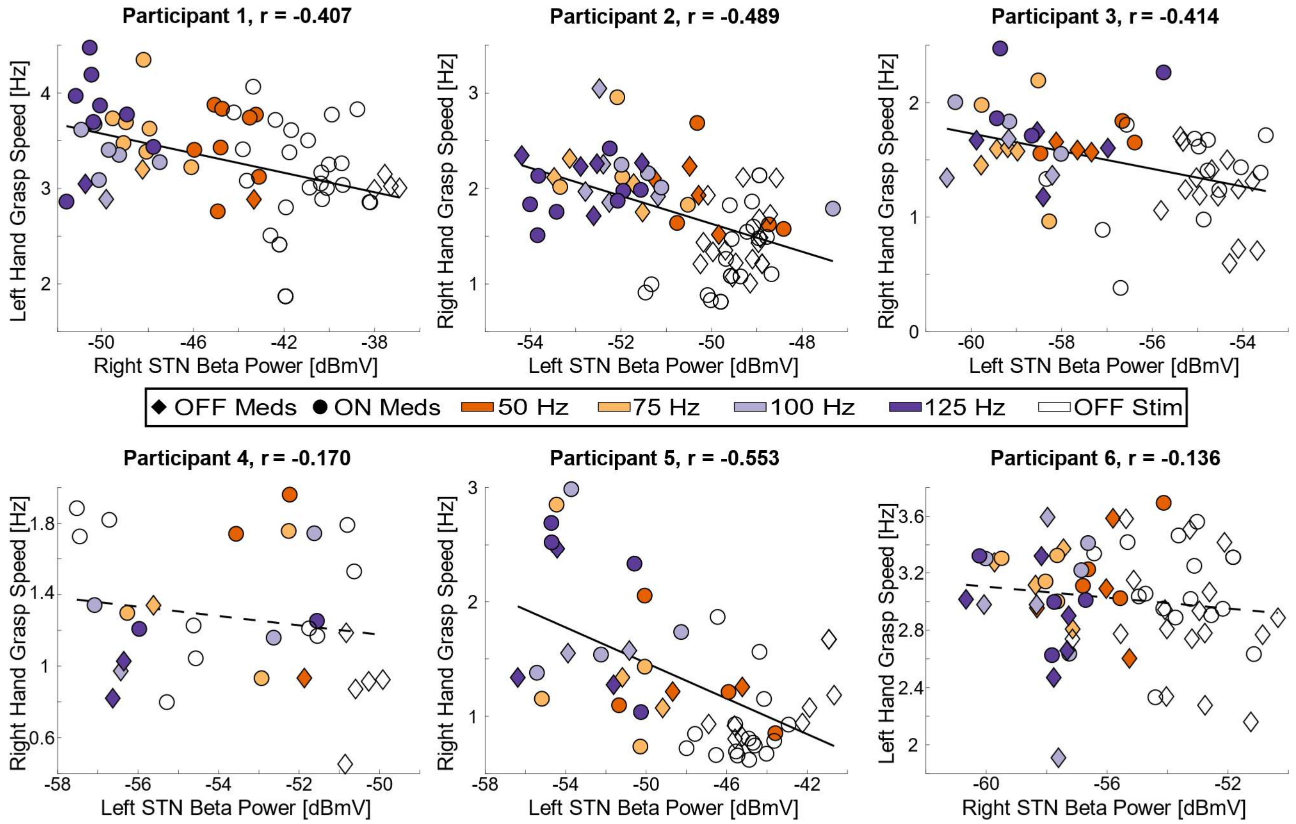


**Figure S6. Best hand grasp speed versus contralateral STN beta power.** For all participants, the hand grasp speed versus beta power in the contralateral STN. Only the hand which had a higher correlation between grip speed and STN beta power is presented. For Participants 1-3 and 5 (and not Participants 4 and 6), there was a significant correlation between bradykinesia and beta power. For significant correlations, best fit lines are depicted in solid black. Dashed lines represent the best linear fit for non-significant correlations.

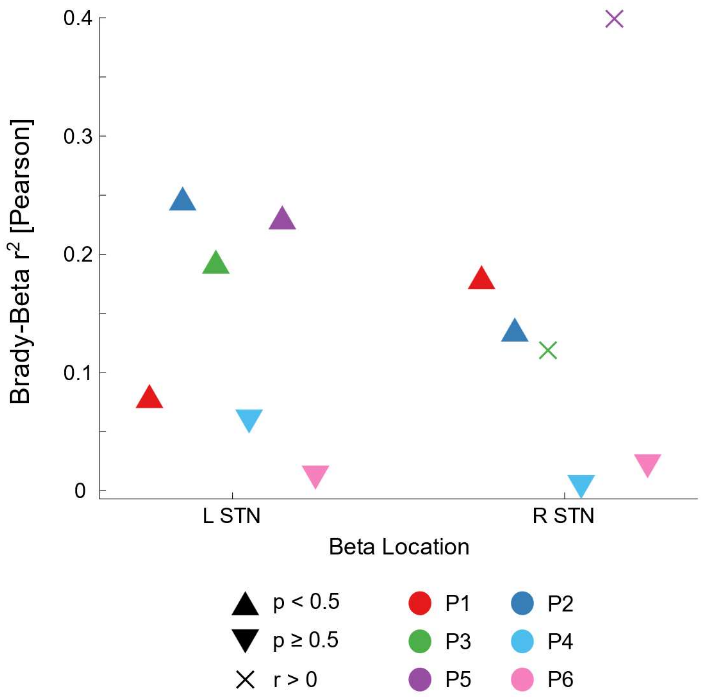


**Figure S7. Correlation between bradykinesia and beta power in the contralateral STN.** Per participant, the Pearson correlation between bradykinesia (i.e. hand grasp speed) and power of the beta oscillations in the contralateral STN. For Participants 1 – 3 and Participant 5, at least one side had a significant anti-correlation. Positive correlations (indicated by X's) were assumed to be caused by DBS artifacts.

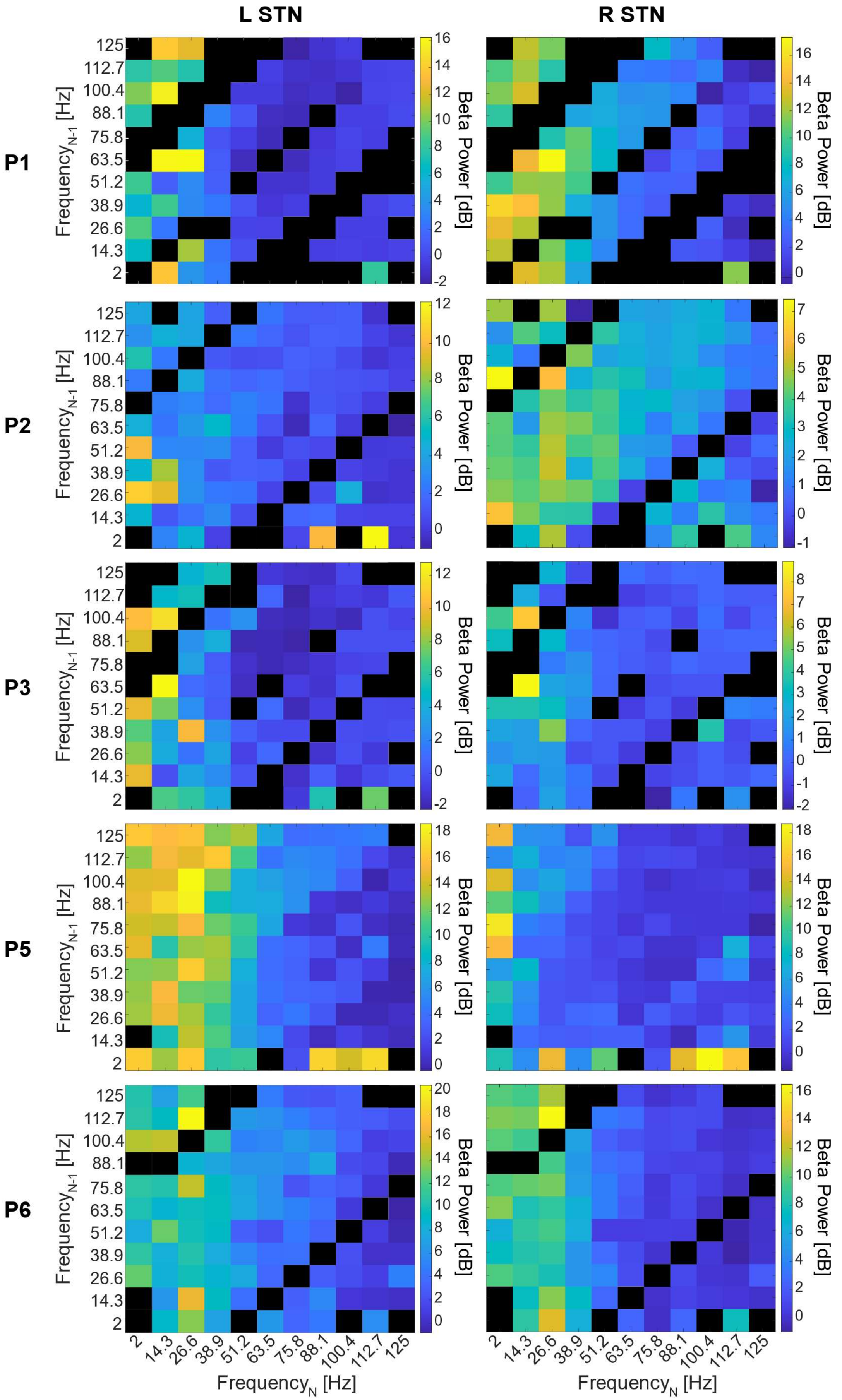
L STN
R STN
P1
P2
P3
P5
P6
Frequency$_{N-1}$ [Hz]
125
112.7
100.4
88.1
75.8
63.5
51.2
38.9
26.6
14.3
2
Beta Power [dB]
Frequency$_N$ [Hz]

**Figure S8. Beta power response relative to the previous frequency.** The mean normalized beta power at each DBS frequency relative to the previous frequency stimulated, for DT DBS random frequency experiments. In general, beta power decreased with increasing frequency of the current DBS frequency (across the horizontal axis), rather than the previous DBS frequency used (vertical). Black squares indicate transitions that did not occur during the experiments.

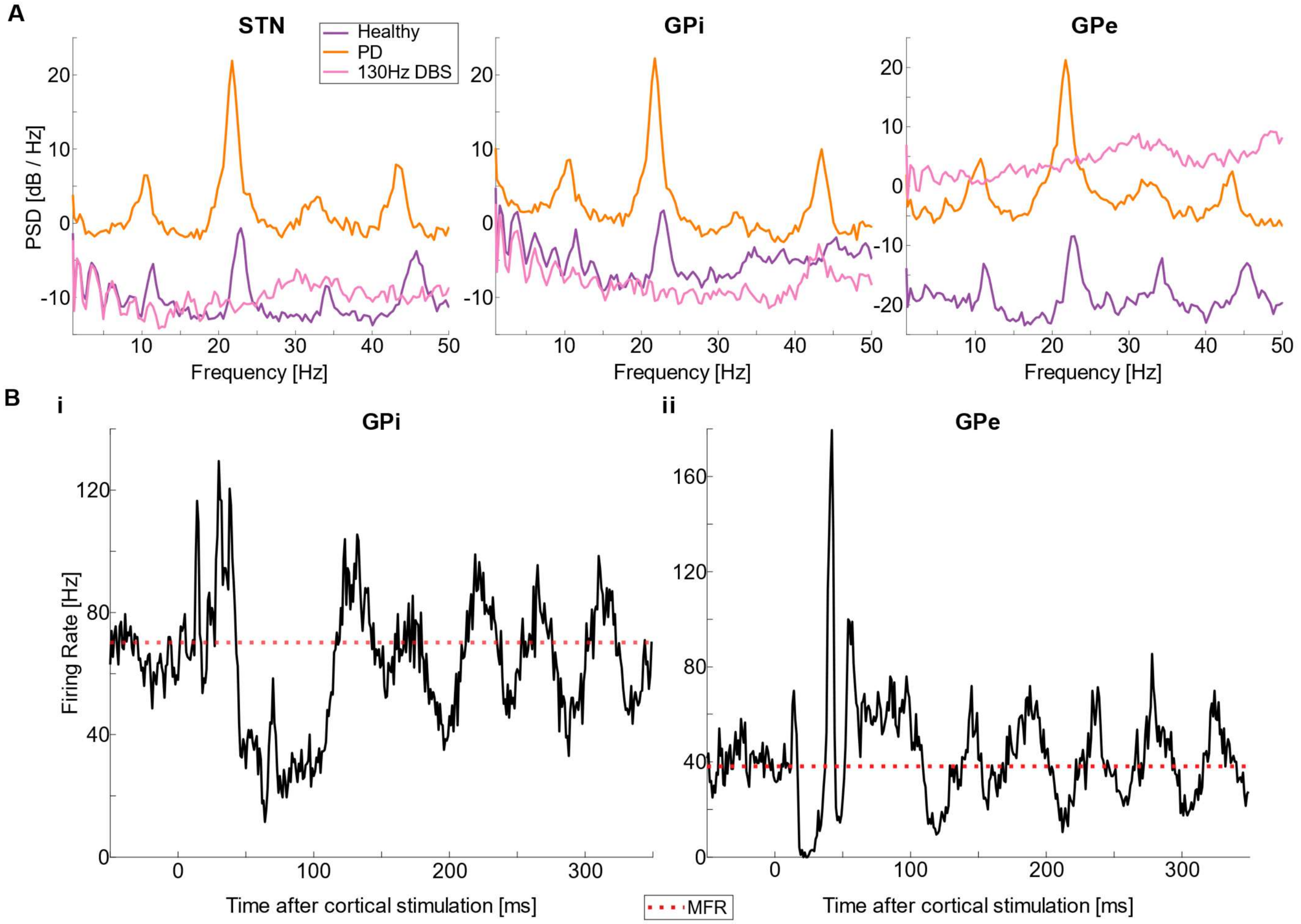


**Figure S9. Model Validation. A**) PSD comparisons of the healthy, Parkinsonian, and 130 Hz STN DBS applied to the Parkinsonian state in the STN, GPi and GPe (left to right). For the Parkinsonian state, there was an increase in power within the beta range compared to the healthy state, with a peak at 22 Hz in all three regions. This peak in the beta band was completely removed by 130 Hz STN DBS. **B)** PSTHs (bin size = 1 ms) of the average of 10 neurons across 100 simulations in the GPi and the GPe (left, right) after stimulation in the cortex. MFR for each region is shown as a red dotted line. Cortical stimulation consisted of one pulse (at t = 0), with a pulse width of 1 ms, in the Parkinsonian state, without DBS. For the GPi (i) there was a peak in activity after the cortical stimulation (0 – 50 ms), followed by a protracted inhibition of activity (50 – 125 ms). Post inhibition, activity peaked (125 – 150 ms) before resuming to normal behavior (~ 200 ms). For the GPe (ii), there was a short inhibition in the activity (10 – 40 ms), followed by a period of excitation in which the GPe reached peak activity (~50 – 100 ms), before resuming baseline activity.

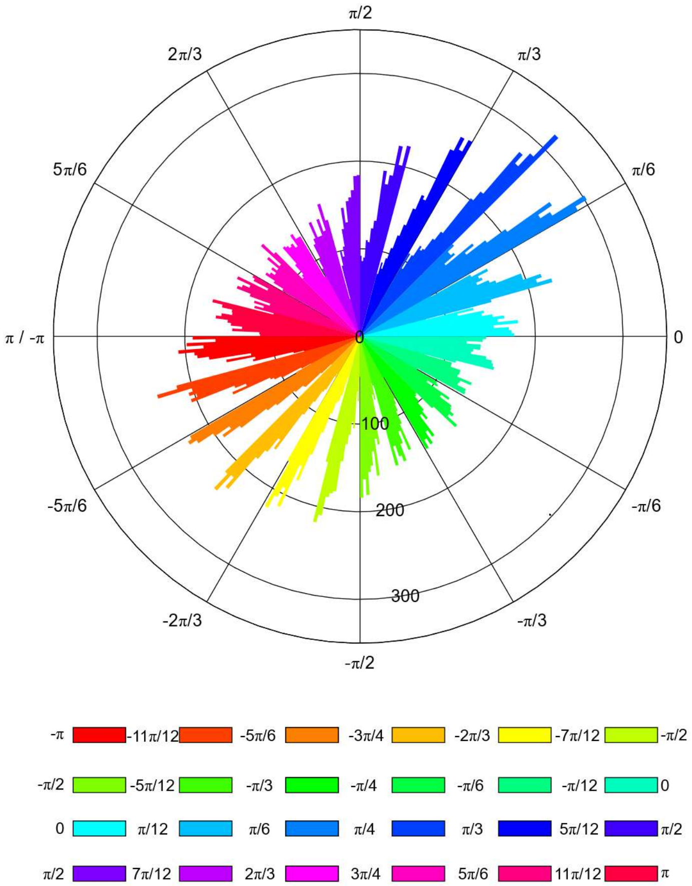


**Figure S10. Controller Accuracy.** For the closed-loop stimulation, we plotted the phase of the LFP in the STN each time a DBS pulse was delivered. Histograms were made of the LFP phase of each pulse, each color in the legend representing the trials in which the controller boundaries started (left) and ended (right). For each stimulation setting, DBS pulses were delivered only within marked bounds, confirming the accuracy of the controller.

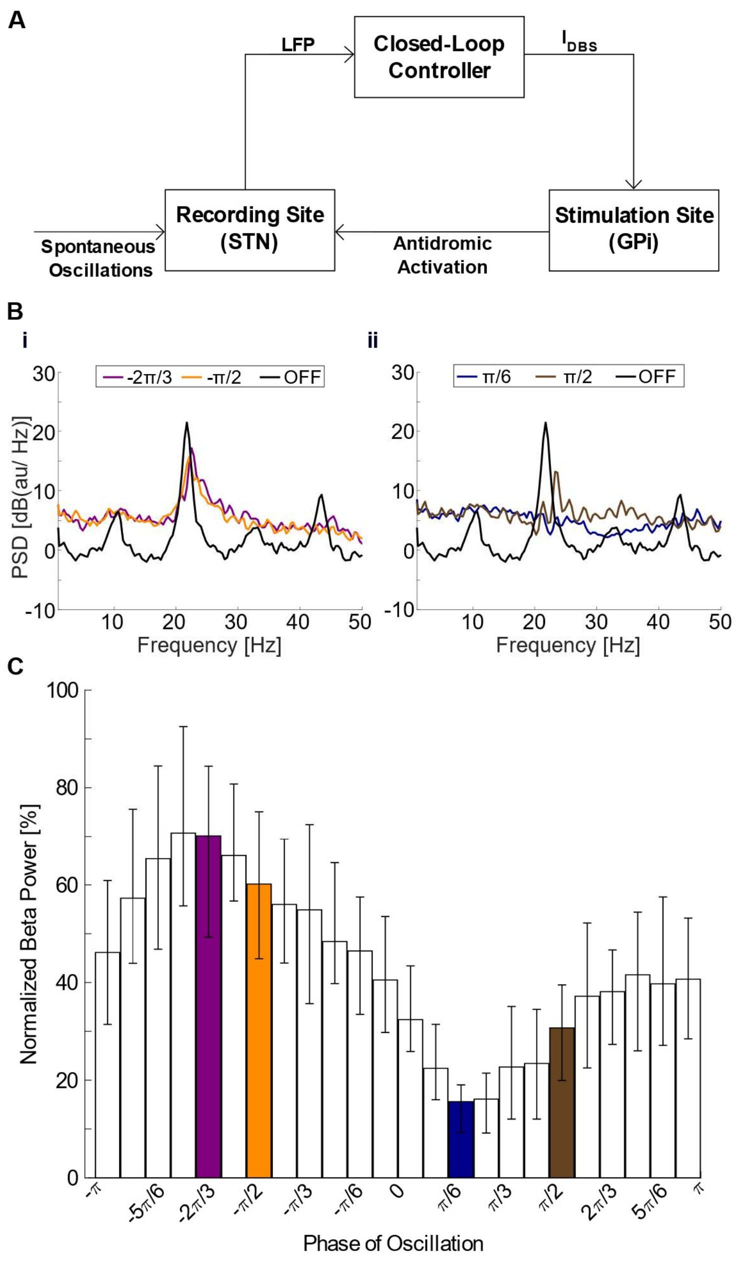


**Figure S11. Effect of GPi DBS delivered at different phases of the ongoing beta oscillations in STN**. **A)** Schematic of the controller. STN LFP was processed to extract the instantaneous phase of the beta oscillations. If the phase was within the epoch, the controller delivered a DBS pulse to the GPi and then deactivated for 20 ms. **B)** PSD plots for different conditions in target phases (colored lines), compared to DBS off PSD (black line). **C)** Normalized beta power across target phase epochs. Error bars show the range of results across ten trials with different random seeds in the model.

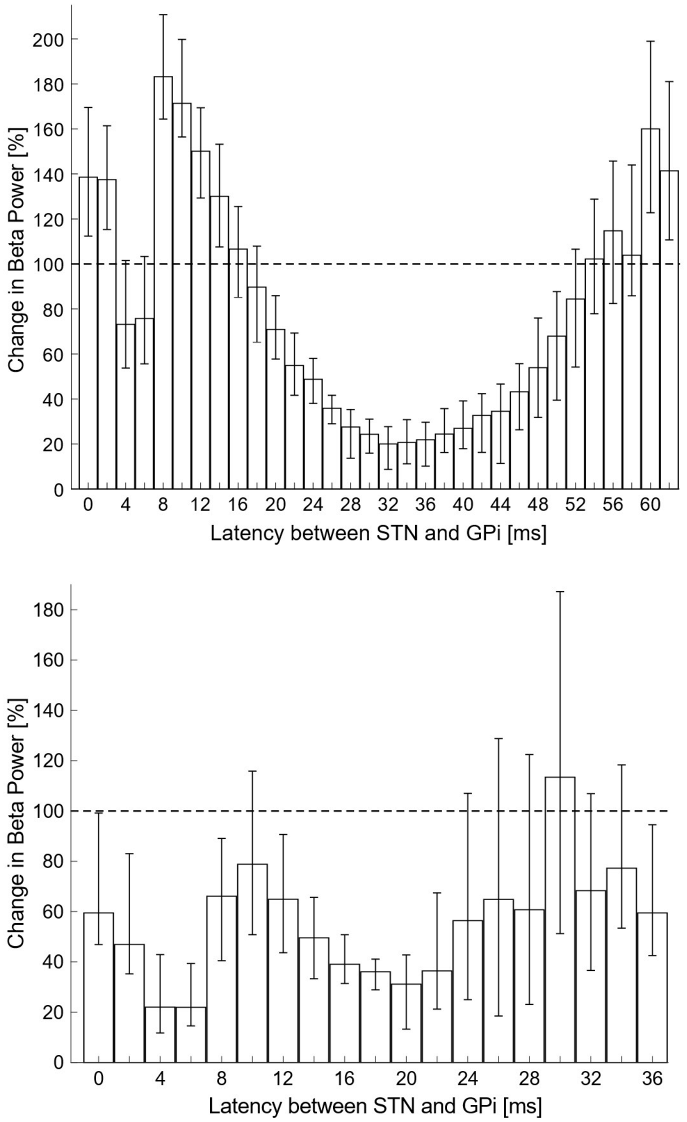


**Figure S12. Latency simulations.** Beta power during DBS across intrahemispheric latencies for 16 Hz (top) and 28 Hz (bottom) DT DBS. The latency range was restricted to the length of the DBS frequency. From these results, we can observe that even if the DBS frequency does not exactly match the frequency of peak power of the beta oscillations (i.e. 22 Hz on the model), a residual beta power of 20.0 % (at 16 Hz DBS) and 22.0% (at 28 Hz DBS) can be observed by making changes in the latency. However, the latency that best-reduced beta power depended on DBS frequency. In both plots, beta power during the DBS off state was at 100% and signaled by a dotted line. Error bars represent the range of beta power at a given latency across 10 trials.

#### Supplemental Tables

**Table S1.** Best (most correlated) hand and contralateral STN hemisphere as determined by the cDBS experiments.

| | Best Hand | Best STN |
|---|---|---|
| Participant 1 | Left | Right |
| Participant 2 | Right | Left |
| Participant 3 | Right | Left |
| Participant 4 | Right | Left |
| Participant 5 | Right | Left |
| Participant 6 | Left | Right |

**Table S2.** Synaptic delay modifications between rodent and primate models.

| | STN-GPe | STN-GPi | GPe-STN | GPe-GPe |
|---|---|---|---|---|
| Kumaravelu 2016 (Rodent)(ms)(11) | 2 | 1.5 | 4 | 1 |
| Modified Model (Primates)(ms) | 5.5 (4) | 4.7 (4) | 6 (5) | 4 (5) |

**Table S3.** Changes to the bias current between the rodent and primate models.

| | GPi | GPe | Thalamus | STN |
|---|---|---|---|---|
| Kumaravelu 2016 (μA/cm$^2$)(11) | 3 | 3 | 1.2 | 0 |
| Modified Model (μA/cm$^2$) | 6 | 6 | 4 | 4 |

**Table S4.** Comparison of model-based mean firing rates with experimental firing rates obtained from the literature.

| MFR | STN | GPi | GPe |
|---|---|---|---|
| Experiment (PD, Primates) | 50.3 (6) | 70.4 (7) | 40.5 (7) |
| Model (PD, Rodents)(11) | 16.9 | 40.8 | 28.75 |
| Modified Model (PD, Primates) | 49.80 | 70.2 | 38.10 |

**Table S5.** Random intercept model to assess the association of stimulation frequency with grip speed (DBS experiments)

| **Effect** | **Estimate (95% CI)** | **P-values** |
|---|---|---|
| Stimulation Frequency: No stimulation | Reference | Reference |
| Stimulation Frequency: 50 Hz | 0.35 (0.20, 0.50) | <0.0001 |
| Stimulation Frequency: 75 Hz | 0.43 (0.28, 0.58) | <0.0001 |
| Stimulation Frequency: 100 Hz | 0.41 (0.26, 0.56) | <0.0001 |
| Stimulation Frequency: 125 Hz | 0.48 (0.34, 0.61) | <0.0001 |
| Medication: Off | Reference | Reference |
| Medication: On | 0.13 (0.03, 0.23) | 0.01 |

**Table S6**. Random intercept model to assess the association of stimulation frequency with beta power (random frequency experiments) using data from the best recording side

| **Effect** | **Estimate (95% CI)** | **P-values** |
|---|---|---|
| 10-second stimulation interval | Reference | Reference |
| 2-second stimulation interval | 1.875 (1.751, 1.999) | <.0001 |
| 4-second stimulation interval | 1.254 (1.075, 1.432) | <.0001 |
| DBS frequency 14.3 vs. 2 Hz for ST DBS | -0.589 (-0.710, -0.469) | <.0001 |
| DBS frequency 26.6 vs. 14.3 Hz for ST DBS | -0.791 (-0.881, -0.702) | <.0001 |
| DBS frequency 38.9 vs. 26.6 Hz for ST DBS | -1.196 (-1.243, -1.148) | <.0001 |
| DBS frequency 51.2 vs. 38.9 Hz for ST DBS | -1.634 (-1.719, -1.549) | <.0001 |
| DBS frequency 63.5 vs. 51.2 Hz for ST DBS | -1.432 (-1.503, -1.361) | <.0001 |
| DBS frequency 75.8 vs. 63.5 Hz for ST DBS | -0.646 (-0.717, -0.574) | <.0001 |
| DBS frequency 88.1 vs. 75.8 Hz for ST DBS | -0.171 (-0.256, -0.086) | <.0001 |
| DBS frequency 100.4 vs. 88.1 Hz for ST DBS | -0.139 (-0.186, -0.092) | <.0001 |
| DBS frequency 112.7 vs. 100.4 Hz for ST DBS | -0.179 (-0.267, -0.090) | <.0001 |
| DBS frequency 125.0 vs. 112.7 Hz for ST DBS | -0.199 (-0.319, -0.079) | 0.0011 |
| DBS frequency 14.3 vs. 2 Hz for DT DBS | -0.661 (-0.805, -0.517) | <.0001 |
| DBS frequency 26.6 vs. 14.3 Hz for DT DBS | -0.925 (-1.032, -0.818) | <.0001 |
| DBS frequency 38.9 vs. 26.6 Hz for DT DBS | -1.453 (-1.509, -1.396) | <.0001 |
| DBS frequency 51.2 vs. 38.9 Hz for DT DBS | -2.023 (-2.124, -1.922) | <.0001 |
| DBS frequency 63.5 vs. 51.2 Hz for DT DBS | -1.748 (-1.833, -1.663) | <.0001 |
| DBS frequency 75.8 vs. 63.5 Hz for DT DBS | -0.708 (-0.793, -0.623) | <.0001 |
| DBS frequency 88.1 vs. 75.8 Hz for DT DBS | -0.107 (-0.208, -0.005) | 0.04 |
| DBS frequency 100.4 vs. 88.1 Hz for DT DBS | -0.113 (-0.167, -0.058) | <.0001 |
| DBS frequency 112.7 vs. 100.4 Hz for DT DBS | -0.204 (-0.306, -0.102) | <.0001 |
| DBS frequency 125.0 vs. 112.7 Hz for DT DBS | -0.250 (-0.388, -0.112) | 0.0004 |
